# Global COVID-19 vaccination uptake across the emergency and early post-emergency periods, 2021-2024

**DOI:** 10.64898/2026.07.02.26356844

**Authors:** Donald J. Brooks, Ashley Germann, Laura Craw, Laure Dumolard, Yoann Nedelec, Ayşe Açma, Craig Schultz, Franck Mboussou, Ajiri Atagbaza, Reena H. Doshi, Marcela Contreras, Martha Velandia Gonzalez, Rafael Léon, Osama Mere, John Kissa, Roberta Pastore, Tondo Opute Emmanuel Njambe, Glenda Gonzales, Benjamin Bayutas, Diana Chang Blanc, Sophie Von Dobschütz, Annelies Wilder-Smith, Marta Gacic-Dobo, Maria D. Van Kerkhove, Katherine L. O’Brien

## Abstract

COVID-19 vaccination helped change the course of the pandemic and remains critical for protecting high-risk groups. We analyzed data submitted to WHO to describe COVID-19 vaccination coverage during the emergency period from 2021-2023 and annual uptake in 2024, the first post-emergency calendar year after the Public Health Emergency of International Concern designation was lifted. By end-2023, global complete primary series coverage reached 65% in the total population, 82% among older adults, and 91% among health and care workers (HCWs), across reporting countries. Booster coverage was lower, at 33%, 60%, and 69%, respectively. Across indicators, disparities by country income group and region emerged early and persisted through 2024. In 2024, vaccination of older adults and HCWs was limited and heterogeneous. These findings underscore the need for stronger, sustainably financed adult immunization platforms and associated monitoring systems to enhance life-course vaccination benefits and to support future outbreak, epidemic, and pandemic responses.

## Introduction

Coronavirus disease 2019 (COVID-19) has had profound effects on population health, health systems, and societies globally since its emergence in late 2019. The COVID-19 pandemic is estimated to have caused 22.1 million excess deaths from 2020-2023^1^. Further, estimates suggest approximately 145 million people worldwide developed post-COVID-19 condition (PCC; also known as long COVID) from 2020-2021, representing a substantial global burden of post-acute morbidity^2^. Beyond its direct health impacts, the pandemic caused major economic and social disruption, with projected cumulative economic output losses over the 2020-2025 period estimated at US$22 trillion relative to pre-pandemic projections^3,4^. Although the risk of severe COVID-19 has diminished with increasing population immunity and improved clinical management, SARS-CoV-2 continues to circulate widely, and both acute COVID-19 and PCC remain ongoing public health concerns^5,6^.

COVID-19 vaccination has been a critical tool for reducing severe disease, death, and longer-term sequelae following SARS-CoV-2 infection, particularly among high-risk groups. During the acute phase of the pandemic, when much of the world’s population was immunologically naive and transmission was at its highest, vaccination helped alter the course of the pandemic and mitigate its health, economic, and social impacts. These benefits were not experienced equally, however. Early country access to vaccines was uneven, and differences in supply, policy, delivery capacity, and demand contributed to marked heterogeneity in country uptake and impact across settings, even as global and regional mechanisms, including COVAX, supported vaccine access and deployment^7–9^. Global modelling studies estimate that COVID-19 vaccination prevented over 20 million deaths during the first year of the rollout, although these benefits were concentrated where access and uptake were achieved earlier and at scale^10,11^.

As COVID-19 management has shifted from emergency response to longer-term control, vaccination remains a critical tool, though programme priorities and challenges have changed^12^. During the emergency period, the dominant task was the rapid, prioritized scale-up of primary vaccination at the population level^13^. In the post-emergency period, emphasis has shifted to sustaining protection among groups at highest risk of severe outcomes, including older adults, persons with chronic conditions, and other clinically vulnerable populations^14^. This has required periodic revaccination, further reinforced in some settings by updated vaccine formulations, in the context of waning immunity and continued viral evolution^15,16^. This change has taken place as countries have moved away from exceptional emergency delivery of COVID-19 vaccination and towards its integration into routine immunization services. Sustained uptake now depends on the ability of these delivery platforms to repeatedly reach high-risk groups over time^17^.

In this context, monitoring COVID-19 vaccination uptake provides core data that support efforts to optimize vaccination programmes and strengthen their impact. As policies shift towards periodic revaccination of high-risk groups and delivery increasingly occurs through routine primary healthcare platforms, uptake monitoring is needed to assess programme reach, identify persisting gaps, and inform programmatic action and potential adjustments. This is especially important because, although most populations now have some degree of immunity from prior infection, vaccination, or both, protection is heterogeneous, wanes over time, and may be affected by viral evolution. With SARS-CoV-2-specific surveillance and direct immunologic monitoring limited in many settings, COVID-19 vaccination uptake therefore remains an important indicator of recent vaccine-derived protection among high-risk groups^5^.

This study presents a global longitudinal analysis of COVID-19 vaccination coverage during the emergency period from 2021-2023 and of reported vaccination uptake in 2024, the first post-emergency calendar year after the Public Health Emergency of International Concern (PHEIC) designation was lifted. From 2021-2023, the World Health Organization (WHO) produced real-time COVID-19 vaccination coverage estimates and disseminated them through multiple monitoring products^18,19^. Building on these efforts, this report provides, to our knowledge, the first globally harmonized time series analysis of COVID-19 vaccination coverage across the emergency period, including among older adults and health and care workers (HCWs), and the first empirical description of reported global uptake in 2024. We aim to characterize the pace and extent of COVID-19 vaccination during the emergency period, assess how coverage varied across countries and population groups, and describe how COVID-19 vaccination patterns evolved during the transition to routine programme implementation. Together, these analyses provide a global reference point for understanding the trajectory of the COVID-19 vaccination response and a baseline for interpreting future uptake trends.

## Methods

### Ethics statement

This study used aggregate, country-reported administrative vaccination data submitted to WHO and did not include individual-level personal data. Ethics approval and informed consent were not required.

### Data sources and definitions

#### Vaccine administration, uptake, and policy data

Countries, areas, and territories (hereafter countries) were advised to monitor COVID-19 vaccination uptake using standardized indicators, as recommended in WHO monitoring guidance^20^. Countries were requested to report a subset of these indicators to WHO through global and regional reporting mechanisms. For 2021-2023, four cumulative indicators were requested: (i) total COVID-19 vaccine doses administered, and the number of individuals with (ii) at least one dose, (iii) a complete primary series (CPS), and (iv) at least one booster dose of a COVID-19 vaccine. For 2024, global monitoring recommendations and reporting requests were revised to a single annual indicator: individuals having received a COVID-19 vaccine dose^21^. Accordingly, 2021-2023 indicators were analyzed as cumulative coverage measures and the 2024 indicator as annual uptake. Standardized indicator definitions are provided in Supplementary Table S1.

WHO guidance recommended monitoring uptake disaggregated by population group prioritized for vaccination (priority group), including older adults and HCWs^20^. Countries reported priority group data using WHO-recommended definitions where possible, although reported data often reflected country-defined definitions. Standardized priority group definitions are summarized in Supplementary Table S2, and country-specific older adult definitions are documented in the accompanying dataset.

COVID-19 vaccination data, including on vaccine administration, uptake, policy, and programme introduction date, were compiled from WHO regional reporting processes, the WHO-UNICEF electronic Joint Reporting Form (eJRF) COVID-19 module, the annual routine immunization eJRF exercise, and, for most European Union Member States in 2024, the European Surveillance System. Reporting mechanisms, reporting frequency, and data compilation processes are described in Supplementary Methods S1.

#### Population definitions and estimates, and vaccination targets

Total and older adult population denominators were derived from the United Nations World Population Prospects 2024 revision by summing age- and sex-specific estimates as required. Analyses for 2021-2023 used 2021 population estimates, and analyses for 2024 used 2024 estimates. Older adult denominators were aligned to country-defined definitions where available; otherwise, older adults were defined as persons aged 60 years and older.

HCW population estimates were obtained from ILOSTAT using the latest available modelled estimates (2019) for the health and care sector. For countries without available ILOSTAT estimates, denominators were derived from pooled estimates among countries in the same WHO region and income group, following previously described WHO methods^22^.

HCW vaccination targets, defined as the number of HCWs a country aimed to vaccinate, were reported by countries alongside uptake data and were often used by countries as operational denominator surrogates where comprehensive HCW population denominators were not available.

#### Country classifications

Countries were classified by World Bank income group (2025 revision), WHO region (2025 affiliations), and COVAX Facility participation category (December 2023 classification). The COVAX Facility classification distinguished self-financing countries from those eligible for donor-supported vaccine access through the COVAX Advance Market Commitment (AMC).

#### Analytic periods

Two analytic periods were defined. January 2021 to December 2023 was defined as the emergency period, reflecting the period of rapid COVID-19 vaccine deployment and scale-up and largely coinciding with the COVID-19 PHEIC. This period was defined by calendar year to align with the structure of WHO vaccination reporting and to provide end-2023 coverage estimates. Although vaccination began in some countries in late 2020, pre-2021 reports in the WHO administrative datasets were sparse and were not included as separate time points in the primary time series analyses. Because uptake indicators were reported cumulatively, vaccinations administered before January 2021 were retained in subsequent cumulative totals and were therefore not excluded from endpoint coverage estimates. January to December 2024 was defined as the first post-emergency calendar year, corresponding to the first full calendar year after the PHEIC designation was lifted.

### Data cleaning and adjustment

#### Data cleaning

An anomaly detection and correction framework was applied to cumulative uptake data for 2021-2023 to identify implausible reporting artefacts and construct monotonic monthly time series. Flagged observations were set to missing, internal missing values were interpolated, and the latest available cumulative value was carried forward after the final report. For total doses administered, at least one dose, and CPS indicators, a further interpolation step was used to construct trajectories between programme introduction and the first reported cumulative value. For older adult and HCW booster indicators, interpolation began at the country’s first reported total population booster value and extended to the first population group-specific report. Total population booster values were not interpolated before the first report. Further details are provided in Supplementary Methods S2 and will be reported separately.

The anomaly detection and correction framework was not applied to 2024 data because the analysis used one annual uptake value per country and priority group, rather than a cumulative time series. For each country, annual and quarterly source extracts were reconciled to derive a single end-2024 uptake value for older adults and HCWs. Where an annual 2024 report was available, that value was used. Where no annual report was available, the latest available quarterly cumulative value for 2024 was used as the end-2024 value. Non-response codes and other missing-value codes were treated as missing, while reported zero values were retained as true reported values. Data review decisions on source prioritization, reporting basis, exclusions, and replacements were documented in an audit log, included in the accompanying dataset, and applied before deriving the final 2024 uptake value.

#### Data adjustment

Following data cleaning, reported numbers of vaccinated HCWs for 2021-2023 were adjusted using previously described WHO methods to account for cases in which reported HCW vaccination targets were lower than estimated HCW population denominators^22^. For countries reporting an HCW vaccination target lower than the ILOSTAT HCW population denominator, the corresponding total population coverage estimate at the same time point was applied to the difference between the HCW population denominator and the reported HCW vaccination target. This estimated additional number of vaccinated HCWs was then added to the reported HCW count. The adjustment was applied separately to each vaccination indicator. No adjustment was applied where no HCW vaccination target was reported or where the target equaled or exceeded the HCW population denominator. Adjusted HCW counts were constrained not to exceed the HCW population denominator. For 2024, HCW estimates were based on direct country-reported numbers of vaccinated HCWs, without the redistribution adjustment applied to 2021-2023 data; coverage estimates were capped at 100%.

Counts for the total population and older adults were constrained not to exceed the corresponding population denominator; no other adjustments were applied.

### Vaccination coverage and uptake analysis

Analyses were limited to the 194 WHO Member States, as of 2025. Analyses disaggregated by COVAX Facility participation category additionally included Kosovo and occupied Palestinian territory, including east Jerusalem, yielding 196 reporting entities.

For 2021-2023, weekly and monthly reported uptake data were harmonized to monthly time series by selecting the latest reported cumulative value within each calendar month. End-2023 coverage estimates were defined as the latest values available as of December 2023. For 2024, uptake was assessed using the reconciled annual country-level values described above.

Patterns of reporting completeness were assessed; methods for this analysis are provided in Supplementary Methods S3.

#### Country-level COVID-19 vaccination coverage estimation

Using cleaned, adjusted uptake and population estimates, coverage and uptake were calculated at the country-level for each indicator and population group. For 2021-2023, coverage estimates were calculated for the total population, older adults, and HCWs. For 2024, uptake estimates were calculated for older adults and HCWs only; total population uptake was not estimated, reflecting changes to global COVID-19 vaccination recommendations^14,21^. Coverage and uptake were estimated only where both numerator and denominator data were available.

#### Aggregate, country grouping COVID-19 vaccination coverage and uptake estimation

Population-weighted aggregate coverage and uptake estimates were calculated by summing cleaned, adjusted uptake figures across countries and dividing by the corresponding summed population estimates, overall and by WHO region, World Bank income group, and COVAX Facility participation category, and policy recommendation category, where relevant. For emergency period time series analyses, denominators were held constant within each grouping, defined as the summed population of countries reporting at least one value for the relevant indicator and population group by December 2023. Countries with missing coverage or uptake estimates were excluded from the relevant aggregate calculations, and countries without an assigned income group were excluded from income group-specific analyses.

Cross-country variation was summarized using the median and interquartile range (IQR; 25th-75th percentiles) of country-level coverage. For emergency-period time series analyses, monthly statistics were presented only when reporting countries in that month represented at least 10% of countries that reported data for the relevant population group and indicator.

#### Coverage parity analysis between priority groups and total population

Country-level coverage in older adults and HCWs was compared with total population coverage at end-2023. Parity was defined as equal coverage in the priority group and total population. Departures from parity were summarized using a coverage ratio and an absolute percentage point difference. Supplementary analyses compare parity measures between end-2021 and end-2023.

### Daily vaccination rate analysis

Estimated daily vaccination rates were reconstructed from weekly country-reported cumulative total dose administration records. Records with missing country identifiers, missing cumulative dose counts, negative cumulative dose counts, or dates outside 1 January 2021 to 31 December 2023 were excluded. Where duplicate records remained for the same country and reporting date, the largest cumulative total dose value was retained. Cumulative total dose series were then cleaned for monotonicity using the same anomaly detection framework as the monthly coverage analyses.

For each country, a daily cumulative series was constructed from the programme introduction date to the last reported cumulative value. Where the programme introduction date was unavailable or did not precede the first report, the day before the first report was used as the zero anchor. Daily cumulative totals between reported dates were linearly interpolated. Daily doses administered were calculated as the day-to-day change in the interpolated cumulative series, and daily vaccination rates were summarized as a trailing 28-day mean. Rates were expressed as doses administered per day and, where relevant, as daily doses administered per 100 population.

Population-weighted aggregate rates were calculated globally and by WHO region, World Bank income group, and COVAX Facility participation category using the same constant denominator approach as the coverage time series analyses. Peak rates were defined as the maximum population-weighted trailing 28-day mean during 2021-2023. Cross-country variation at each time point was summarized using the unweighted 10th-90th percentile range across reporting countries.

### Vaccination policy analysis

National COVID-19 vaccination policies for 2024 were classified as (i) primary vaccination and periodic revaccination recommended, (ii) primary vaccination recommended, or (iii) vaccination not recommended. Overall summaries excluded countries with missing responses and used reporting WHO Member States as the denominator. Income group-stratified summaries included all WHO Member States with an income group classification, with missing responses retained as a separate “No response provided” category. Results are presented as counts and percentages of reporting WHO Member States, stratified by population group and World Bank income group.

## Results

### Emergency period (2021-2023)

#### Final cumulative COVID-19 vaccination coverage by end-2023

Globally, population-weighted complete primary series (CPS) coverage reached 65% in the total population, 82% among older adults, and 91% among HCWs by December 2023 (Table 1, Figure 1a). In contrast, coverage with at least one booster dose remained substantially lower across all population groups, reaching 33% in the total population, 60% among older adults, and 69% among HCWs, across reporting countries.

**Figure 1.**
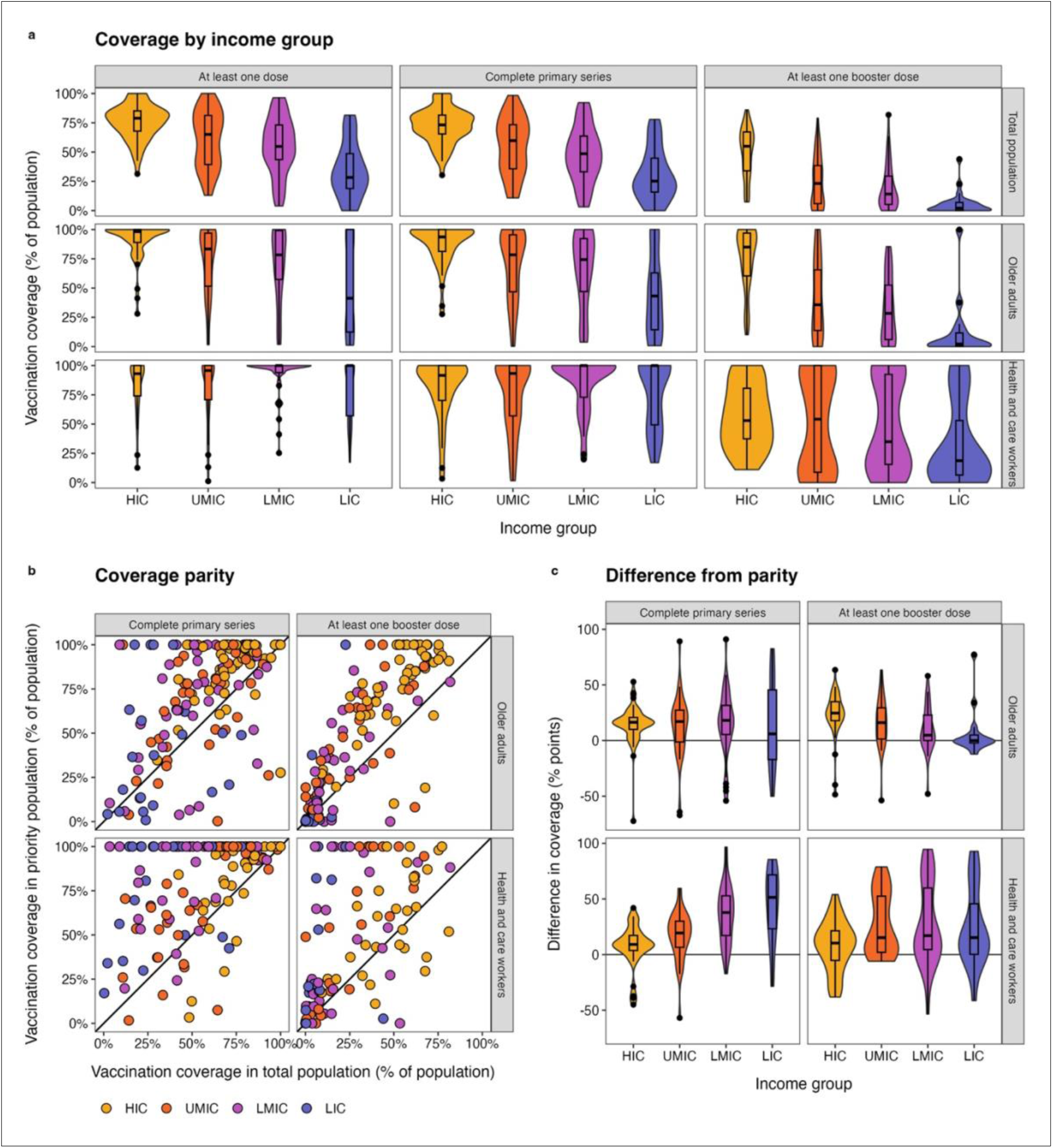
COVID-19 vaccination coverage and priority-group coverage relative to the total population, end-2023. (a) Violin plots show the distribution of country-level vaccination coverage for at least one dose, complete primary series, and at least one booster dose, stratified by population group (total population, older adults, and health and care workers) and World Bank income group. Boxes indicate medians and interquartile ranges; points represent outlier countries. Coverage is expressed as a percentage of the relevant population group. (b) Each point represents a country, with vaccination coverage in the total population shown on the x-axis and vaccination coverage in the priority group shown on the y-axis, separately for complete primary series and at least one booster dose among older adults and health and care workers. The diagonal line indicates parity between priority-group and total-population coverage. (c) Distributions of country-level differences in vaccination coverage between priority groups and the total population (priority group minus total population, percentage points), stratified by World Bank income group. Positive values indicate higher coverage in priority groups. Violin plots show the distribution of country-level estimates, with embedded boxplots indicating medians and interquartile ranges; points denote outlier countries.

**Table 1.** COVID-19 vaccination coverage by end-2023, by population group, income group, WHO region, and COVAX Facility participation category. Values shown are population-weighted aggregate coverage estimates, expressed as a percentage of the relevant population group. Coverage is shown separately for complete primary series and at least one booster dose among the total population, older adults, and health and care workers, overall and by World Bank income group, WHO region, and COVAX Facility participation category. *The number of countries reporting vaccination coverage varied by population group and dose; reporting completeness is visualized in Supplementary Figure S1*. **Note:** CPS = Coverage with a complete primary series; booster = coverage with at least one booster dose

| Category | Total population |  | Older adults |  | Health and care workers |  |
| --- | --- | --- | --- | --- | --- | --- |
|  | CPS | Booster | CPS | Booster | CPS | Booster |
| **All** |  |  |  |  |  |  |
| Global | 65% | 33% | 82% | 60% | 91% | 69% |
| **Income group** |  |  |  |  |  |  |
| HIC | 72% | 52% | 92% | 82% | 92% | 61% |
| UMIC | 77% | 46% | 82% | 66% | 93% | 79% |
| LMIC | 60% | 19% | 78% | 33% | 88% | 73% |
| LIC | 27% | 4% | 34% | 4% | 67% | 15% |
| **WHO region** |  |  |  |  |  |  |
| AFR | 31% | 6% | 64% | 11% | 66% | 16% |
| AMR | 71% | 42% | 94% | 80% | 98% | 88% |
| EMR | 49% | 18% | 62% | 30% | 80% | 56% |
| EUR | 65% | 48% | 80% | 70% | 73% | 50% |
|  | CPS | Booster | CPS | Booster | CPS | Booster |
| SEAR | 70% | 20% | 82% | 35% | 100% | 100% |
| WPR | 84% | 53% | 84% | 73% | 96% | 91% |
| **COVAX participation category** |  |  |  |  |  |  |
| AMC | 54% | 17% | 69% | 27% | 83% | 64% |
| Self-financing | 82% | 53% | 90% | 78% | 96% | 81% |

Pronounced income-related gradients were evident for both CPS and booster coverage across population groups through end-2023 (Table 1, Figure 1a). In the total population, CPS coverage ranged from 27% in low-income countries (LICs) to 72% in high-income countries (HICs), with corresponding booster coverage figures of 4% and 52%, respectively. Among older adults, CPS coverage was highest in HICs (92%) and remained high in upper-middle-income countries (UMICs) and lower-middle-income countries (LMICs) (82%, 78%) but was markedly lower in LICs (34%). Booster dose coverage among older adults showed a sharper gradient, ranging from 4% in LICs to 82% in HICs. HCWs generally achieved high CPS coverage across income groups (88%, 93%, and 92% in LMICs, UMICs, and HICs, respectively), but coverage was lower in LICs (67%). Booster dose coverage among HCWs was higher than in the total population across all income groups (Table 1).

Disparities in coverage were also observed across WHO regions (Table 1). CPS coverage in the total population ranged from 31% in the African Region (AFR) to 84% in the Western Pacific Region (WPR); these same regions also marked the extremes for booster coverage, ranging from 6% in AFR to 53% in WPR. Among older adults, CPS coverage remained comparatively high across regions, ranging from 62% in the Eastern Mediterranean Region (EMR) to 94% in the Region of the Americas (AMR), but booster dose coverage was lower and more uneven, ranging from 11% in AFR and 30% in EMR to 80% in the AMR. Among HCWs, CPS coverage was high in most regions (including 100% reported in the South-East Asia Region (SEAR)), while booster coverage varied markedly, ranging from 16% in AFR to 100% reported in SEAR.

Coverage also differed substantially by COVAX Facility participation category (Table 1). Countries in the self-financing category reported higher coverage (CPS / booster: 82% / 53% in the total population; 90% / 78% in older adults; 96% / 81% in HCWs) than those eligible for donor-supported vaccine access through the COVAX Advance Market Commitment (AMC) (54% / 17% in the total population; 69% / 27% in older adults; 83% / 64% in HCWs).

The distribution of country-level coverage further illustrates these disparities (Figure 1a). Violin plots show substantial coverage heterogeneity within income groups, most notably for booster doses. This heterogeneity was particularly pronounced in LICs and LMICs, where many countries reported very low booster coverage across population groups, though some achieved substantially higher levels. Heterogeneity was less pronounced for CPS, especially in HICs, where coverage more often clustered at higher levels, particularly among older adults.

CPS coverage was generally higher in older adults than in the total population, with many countries falling above the parity line (Figure 1b; Supplementary Figure S2). Similarly, HCW CPS coverage was often higher than in the total population, with most countries above parity, though with greater variability in LICs and LMICs (Figure 1c). For booster doses, the pattern diverged for older adults. Outside HICs, many countries clustered near parity, indicating that booster uptake was often not substantially higher in older adults than in the total population (Figure 1b). Among HCWs, booster coverage also often exceeded total population coverage, although variability across income groups remained notable (Figure 1c).

#### Temporal evolution of COVID-19 vaccination coverage

Cumulative vaccine administration and coverage trajectories differed substantially across income groups, WHO regions, and COVAX Facility participation categories, with separation emerging early in the rollout and persisting through end-2023 (Figure 2; Supplementary Figures S3-S4). Doses administered per 100 population rose steeply during 2021, but accumulated earlier and to higher levels in HICs, self-financing countries, and several higher-uptake regions, whereas LICs, AMC-supported countries, and some regions, particularly AFR, remained on persistently lower trajectories (Figure 2a). Coverage trajectories likewise differed by population group and dose type, with global population-weighted coverage for at least one dose and CPS increasing rapidly through 2021 to early 2022 and then rising more gradually, with consistently higher levels among older adults and HCWs than in the total population (Figure 2b). Booster dose coverage began later, reflecting later authorization and introduction of booster doses, and increased more slowly before plateauing, remaining well below CPS coverage by end-2023 across all population groups. Median country-level trajectories likewise showed substantial heterogeneity, particularly for booster doses, with broad interquartile ranges (IQR) persisting through 2022-2023 and especially pronounced among priority groups and in lower-income settings (Figure 2c; Supplementary Figure S4).

**Figure 2.**
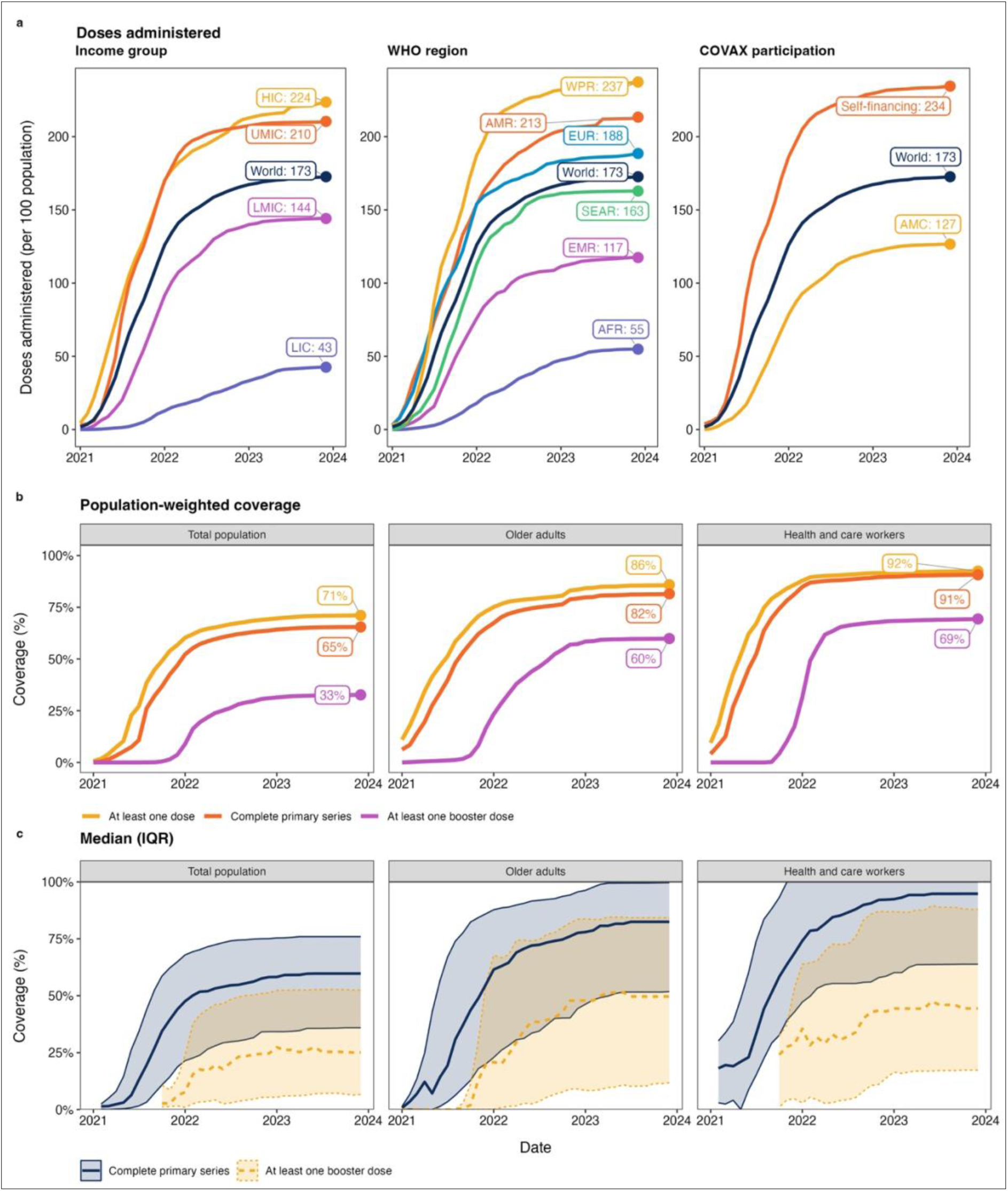
COVID-19 vaccine doses administered and vaccination coverage over time, 2021-2023. (a) Population-weighted cumulative numbers of COVID-19 vaccine doses administered per 100 population in the total population, stratified by World Bank income group, WHO region, and COVAX Facility participation category. (b) Population-weighted global coverage with at least one dose, complete primary series, and at least one booster dose among the total population, older adults, and health and care workers. End-2023 values are labeled. (c) Median country-reported coverage trajectories for complete primary series and at least one booster dose, with shaded bands indicating the interquartile range (25th-75th percentiles) across reporting countries, shown for the same population groups. Estimates are displayed only for months in which reporting countries represented at least 10% of the countries that ever reported that indicator and population group during 2021-2023. Coverage is expressed as a percentage of the relevant population group.

Reporting patterns also changed over time, with implications for interpretation of later-period values (Supplementary Figure S1). Although the cumulative number of Member States that had reported at least once increased over the period for CPS and booster coverage across population groups, the number reporting in any given month declined substantially, with a marked drop-off during 2023, particularly for older adults and HCWs.

#### Dynamics of daily COVID-19 vaccination rates

Daily COVID-19 vaccination rates varied substantially over time and across settings during 2021-2023 (Figure 3; Supplementary Table S3; Supplementary Figure S5). Globally, median daily vaccination rates rose rapidly during 2021, reached a mid-2021 peak, and then declined through 2022 to persistently low levels in 2023 (Figure 3a). The global trajectory showed a bimodal peak pattern, also evident in HICs, likely reflecting the sequencing of first- and second-dose administration. Across income groups, peak activity occurred earlier and at higher intensity in HICs and UMICs than in LMICs and LICs. Population-weighted peak daily vaccination rates ranged from 0.82% of the total population per day in UMICs and 0.64% in HICs to 0.44% in LMICs and 0.12% in LICs, with peaks occurring progressively later from higher- to lower-income settings (Supplementary Table S3).

**Figure 3.**
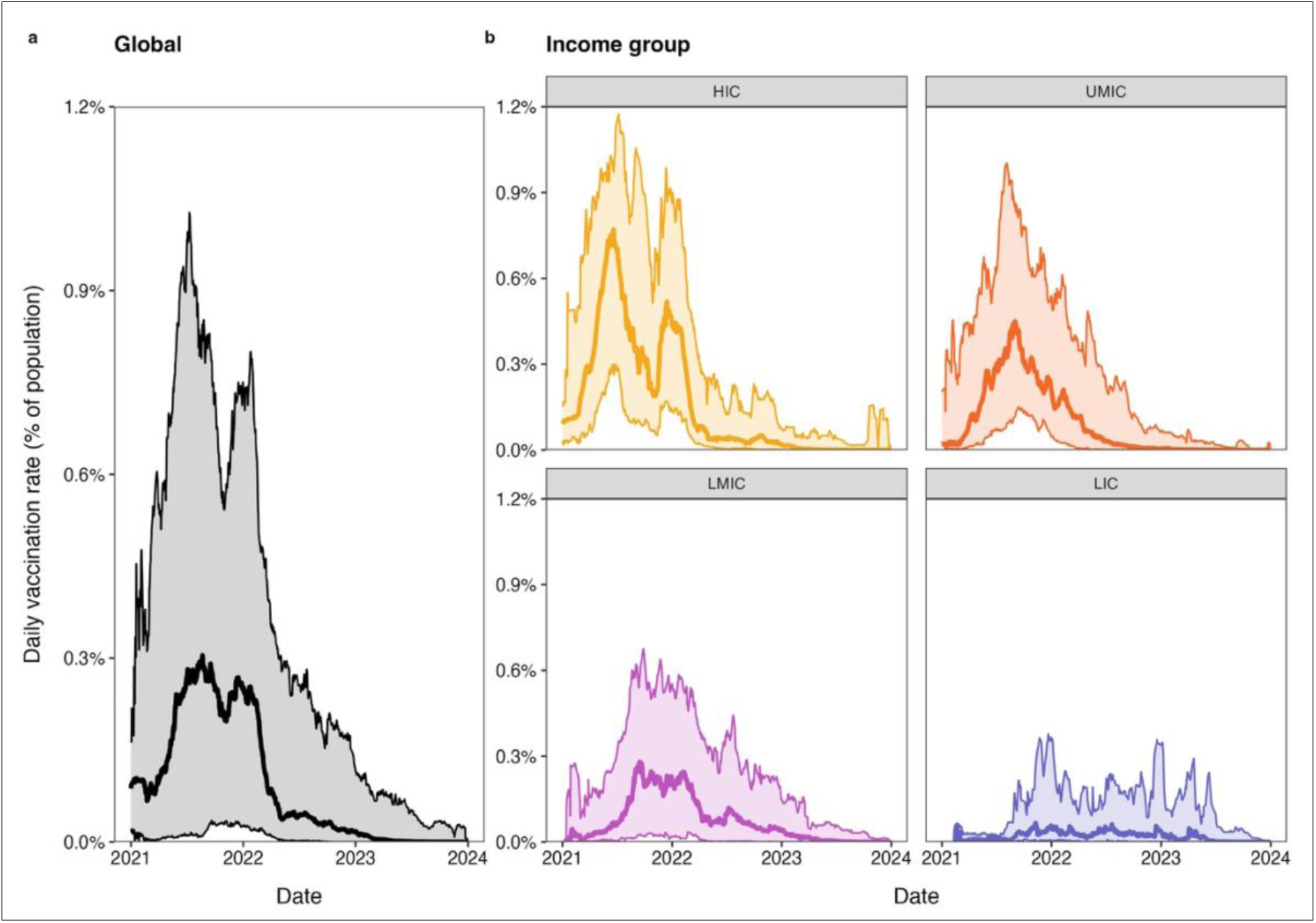
Daily COVID-19 vaccination rate, global and by World Bank income group, 2021-2023. Daily vaccination rates are expressed as a percentage of total population and smoothed using a trailing 28-day mean. Solid lines show the median country-level daily vaccination rate. Shaded areas show the 10th-90th percentile range of country-level rates. (a) Global distribution across WHO Member States. (b) Distributions stratified by World Bank income group.

Substantial within-income group heterogeneity was evident during periods of peak activity, particularly in LMICs and LICs, where median rates were lower overall but intermittent surges persisted over a longer period (Figure 3b). Population-weighted average rates showed related temporal patterns by WHO region, income group, and COVAX Facility participation category, with WPR, HICs, and self-financing countries peaking earlier and at higher intensity than AFR, LICs, and AMC-supported countries (Supplementary Table S3; Supplementary Figure S5). Across groups, daily vaccination rates declined steadily after peak activity and remained low through 2023, reflecting changing vaccination programme goals.

### Early post-emergency period, including integration into routine vaccination programmes (2024)

#### National COVID-19 vaccination policies in 2024

In 2024, national COVID-19 vaccination policies were most commonly focused on priority groups rather than the total population (Figure 4). Among WHO Member States reporting a COVID-19 vaccination policy, recommendations for primary vaccination and periodic revaccination were most common for older adults (108 of 140 reporting countries, 77%), older adults with chronic conditions (99 of 121, 82%), adults with chronic conditions (102 of 126, 81%), and HCWs (90 of 126, 71%) (Figure 4a). For pregnant women, 81 of 123 (66%) reported primary vaccination and periodic revaccination, while 20 of 123 (16%) reported primary vaccination only.

**Figure 4.**
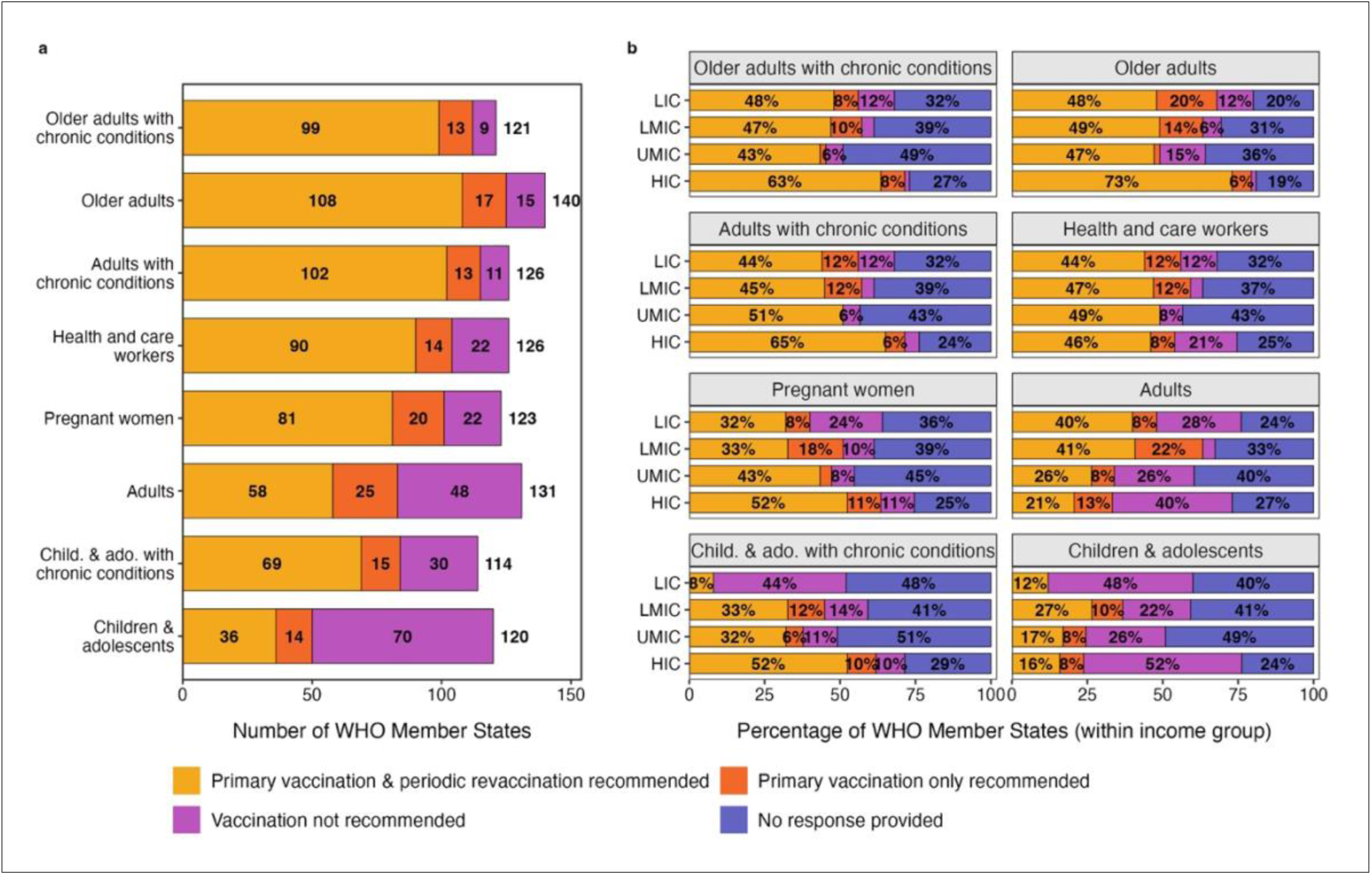
National COVID-19 vaccination policy recommendations for priority groups in 2024. (a) Number of WHO Member States recommending primary vaccination and periodic revaccination, recommending primary vaccination only, or not recommending vaccination for select priority groups in 2024. Values at the end of each bar indicate the total number of reporting Member States for each population group. (b) Percentage distribution of WHO Member States, by World Bank income group, reporting each vaccination policy category for selected priority groups in 2024, including a “no response” category. Percentages are calculated within income group.

National COVID-19 vaccination policies less consistently included broader age-based groups. For adults, 58 of 131 (44%) reported primary vaccination and periodic revaccination, and 48 of 131 (37%) reported that vaccination was not recommended. For children and adolescents, vaccination was most frequently reported as not recommended (70 of 120, 58%), with 36 of 120 (30%) reporting primary vaccination and periodic revaccination. For children and adolescents with chronic conditions, 69 of 114 (61%) reported primary vaccination and periodic revaccination recommendations (Figure 4a).

Substantial variation in policy recommendations and response completeness was observed by income group (Figure 4b), with higher proportions of primary vaccination and periodic revaccination recommendations generally reported in higher-income settings and smallest “no response” shares in HICs across multiple population groups.

#### Annual COVID-19 vaccination uptake in 2024

In 2024, global-level analysis focused on reported uptake among priority groups, consistent with the shift in global vaccination recommendations^14,21^. Globally, 102 WHO Member States reported COVID-19 vaccine uptake among older adults, representing 31% of the global older adult population, with a population-weighted uptake among reporters of 12%. For HCWs, 70 WHO Member States reported uptake, representing 25% of the global HCW population, with a population-weighted uptake among reporters of 6% (Table 2).

**Table 2.** COVID-19 vaccination uptake among priority groups in 2024, by income group, WHO region, and vaccination policy category. Values shown are population-weighted aggregate uptake estimates, defined as the percentage of the relevant population group receiving at least one COVID-19 vaccine dose during 2024. Uptake is shown for older adults and health and care workers, overall and by World Bank income group, WHO region, and vaccination policy category. For each grouping, the table also shows the number of reporting WHO Member States and the proportion of the grouping population represented by reporting countries. **Note:** MS = Member States

| Grouping | Older adults |  |  | Health and care workers |  |  |
| --- | --- | --- | --- | --- | --- | --- |
|  | Count of reporting MS | Proportion of grouping population in reporting MS | Uptake (%) | Count of reporting MS | Proportion of grouping population in reporting MS | Uptake (%) |
| **All** |  |  |  |  |  |  |
| Global | 102 | 31% | 12% | 70 | 25% | 6% |
| **Income group** |  |  |  |  |  |  |
|  | Count of reporting MS | Proportion of grouping population in reporting MS | Uptake (%) | Count of reporting MS | Proportion of grouping population in reporting MS | Uptake (%) |
| HIC | 44 | 51% | 20% | 23 | 22% | 5% |
| UMIC | 26 | 26% | 7% | 23 | 31% | 10% |
| LMIC | 20 | 17% | 5% | 13 | 18% | 0% |
| LIC | 12 | 50% | 10% | 10 | 53% | 2% |
| **WHO region** |  |  |  |  |  |  |
| AFR | 19 | 46% | 1% | 16 | 37% | 1% |
| AMR | 23 | 47% | 13% | 19 | 25% | 15% |
| EMR | 9 | 43% | 8% | 7 | 47% | 2% |
| EUR | 41 | 72% | 16% | 23 | 39% | 4% |
| SEAR | 3 | 3% | 28% | 1 | 0% | 0% |
| WPR | 7 | 12% | 13% | 4 | 10% | 3% |
| **Policy category** |  |  |  |  |  |  |
| Primary vaccination & periodic revaccination recommended | 76 | 62% | 15% | 47 | 51% | 9% |
| Primary vaccination only recommended | 10 | 42% | 3% | 5 | 25% | 2% |
| Vaccination not recommended | 6 | 28% | 0% | 6 | 33% | 2% |
|  | Count of reporting MS | Proportion of grouping population in reporting MS | Uptake (%) | Count of reporting MS | Proportion of grouping population in reporting MS | Uptake (%) |
| No response provided | 10 | 7% | 5% | 12 | 8% | 2% |

Across income groups, uptake among older adults was highest in HICs (20%) and remained lower in UMICs (7%) and LMICs (5%), while LICs reported 10% uptake among older adults. Uptake among HCWs was highest in UMICs (10%), compared with 5% in HICs, and remained low in LICs (2%) and negligible in LMICs (0%) (Table 2). The proportion of the population group represented by reporting countries also varied, ranging from 17% of older adults in LMICs to 51% in HICs, and from 18% of HCWs in LMICs to 53% in LICs.

Marked regional variation was observed. Among older adults, population-weighted uptake ranged from 1% in AFR to 28% in SEAR, with intermediate levels in the European Region (EUR) (16%), AMR (13%), WPR (13%), and EMR (8%). Among HCWs, uptake ranged from 0% in SEAR to 15% in AMR, with 4% in EUR and 1-3% in other regions (Table 2). Regional representativeness also varied substantially, from 3% of older adults in SEAR to 72% in EUR and from 0% for HCWs in SEAR to 47% in EMR.

Uptake also differed by vaccination policy category. Among countries recommending primary vaccination and periodic revaccination, population-weighted uptake was 15% among older adults and 9% among HCWs. Countries reporting uptake data represented 62% and 51% of the respective populations among countries also reporting on vaccination policy. Uptake was lower among reporting countries recommending primary vaccination only, at 3% in older adults and 2% in HCWs, and among countries reporting that vaccination was not recommended, at 0% among older adults and 2% among HCWs (Table 2).

Estimates should be interpreted in the context of more limited reporting than during the emergency period (Table 2).

## Discussion

This study provides the first globally comprehensive assessment of COVID-19 vaccination coverage across both the emergency and early post-emergency periods. It is a harmonized, longitudinal assessment of coverage during the emergency period from 2021-2023, including coverage among older adults and HCWs, and of annual uptake in 2024, the first full calendar year after the PHEIC designation was lifted. By end-2023, global CPS coverage reached 65% in the total population, 82% among older adults, and 91% among HCWs. Further, CPS coverage among older adults and HCWs exceeded that of the total population in most countries, consistent with risk-based prioritization. These findings underscore the broad reach of emergency COVID-19 vaccination efforts and the extent to which priority groups were reached during the primary series rollout, as called for by WHO policy recommendations. At the same time, progress was deeply uneven. Disparities in both the pace and extent of vaccination emerged early, varied considerably across country income groups and regions, and persisted through the end of the emergency period. Beyond the primary series, booster coverage remained substantially lower, reaching only 33% in the total population, indicating that broad initial reach was not matched by sustained or additional vaccination.

This pattern continued into the early post-emergency period. In 2024, annual uptake among priority groups was low and heterogeneous, including in many settings where national policies recommended periodic revaccination. Among countries reporting 2024 uptake data for the respective population group, population-weighted uptake was 12% among older adults and 6% among HCWs, varying substantially across country income groups and regions. During this period, national COVID-19 vaccination policies were most commonly focused on priority groups, in line with WHO SAGE recommendations^14^. Together, findings from the emergency and early post-emergency periods show that rapid scale-up of primary series vaccination did not necessarily translate into sustained uptake of subsequent doses among priority groups. Although COVID-19 vaccination programmes achieved substantial reach in many settings, coverage remained uneven, and 2024 uptake patterns suggest that repeated vaccination of high-risk groups through routine delivery platforms remains a distinct programmatic challenge.

The uneven achievement of primary series coverage during the emergency period reflected inequities in vaccine access, including the early concentration of supply in HICs and delayed availability in many lower-income countries, and countries’ differing capacity to deliver vaccines rapidly at scale. Although these dynamics have been examined extensively elsewhere, our findings add an important temporal dimension^23–26^. Vaccine rollout performance varied not only in scale, meaning the level of coverage ultimately achieved, but also in pace, meaning how quickly this coverage was reached. Daily vaccination rate patterns showed that higher-income settings generally reached earlier and higher delivery rate peaks, whereas lower-income settings scaled up more slowly and at lower peak delivery rates. In the context of a rapidly evolving pandemic, this difference in pace can be consequential. Earlier and faster rollout can avert infections, severe disease, and deaths sooner, while delayed rollout postpones these benefits, even where substantial coverage is eventually achieved^27–29^. These findings point to the importance of assessing pandemic vaccine equity not only by the coverage eventually achieved, but also by how quickly protection reaches populations.

The lower and more heterogeneous uptake observed beyond the primary series suggests that sustaining COVID-19 vaccination became a distinct challenge. This pattern was already apparent in booster dose coverage during the emergency period and became more pronounced in 2024 as vaccination moved into more routine implementation. It likely reflects a combination of shifting epidemiology, changing demand, and evolving programmatic priorities, including increasing population immunity, lower perceived urgency in many settings, and declining prioritization of COVID-19 vaccination within broader health system planning. Although the balance of these factors likely varied across countries, the overall pattern indicates that the conditions that enabled rapid emergency deployment changed over time in ways that affected additional vaccination efforts.

Sustaining uptake was further complicated by the practical difficulty of repeatedly reaching adult risk groups with COVID-19 vaccines. During the emergency period, stand-alone and campaign-based delivery enabled rapid vaccination of adult populations at a scale not previously achieved through routine adult vaccination systems in many settings. As the emergency response receded, these approaches became harder to sustain, with many countries lacking established platforms or financing for periodic revaccination of high-risk adult groups. In response, WHO and UNICEF recommended integrating COVID-19 vaccination into primary health care (PHC) and other existing health delivery platforms, including seasonal influenza vaccination programmes where they exist, to support sustainability^17^. However, the feasibility of integration, and the sustainability gains it can deliver, vary across settings. Although examples of integration are widely documented, published experience suggests that it nevertheless remains operationally challenging where coordination, financing, and adult-focused health service delivery platforms are weak^30,31^.

These findings have important implications for both future outbreak, epidemic, and pandemic vaccine deployments and adult vaccination programmes, including for COVID-19. They point to two critical dimensions of vaccine equity. The first is equitable access to pandemic-related products, a central global preparedness challenge reflected in the WHO Pandemic Agreement, adopted in May 2025, and in continuing negotiations on a Pathogen Access and Benefit Sharing system^32^. The persistent disparities observed during COVID-19 vaccine roll-out underscore the importance of mechanisms that improve timely and equitable access to vaccines and other medical countermeasures during future health emergencies. The second is the capacity to turn access into protection. Once vaccines are available and recommended, countries need operational capacity to identify priority groups, deliver vaccination, monitor uptake, and adapt programmes as the disease epidemiology changes. The emergency period showed the importance of this capacity for rapid vaccine deployment, while the early post-emergency period showed its importance for sustaining vaccination of priority groups over time. Stronger adult immunization delivery platforms, embedded in PHC programmes and aligned with the life-course vaccination approach, are therefore important not only for COVID-19 vaccination and other maternal and adult vaccines, but also for future health emergency responses.

This analysis has several limitations. First, limitations inherent to the source data affected estimation and comparability. Estimation of vaccination coverage and uptake among older adults and HCWs was constrained by the availability and comparability of population denominators; definitions varied across countries, and standard denominators may not have fully aligned with the population groups targeted or reported by countries. Some denominator estimates were incomplete, outdated, or modelled. As a result, some coverage and uptake estimates may reflect differences between reported numerator definitions and denominator definitions, rather than true differences in vaccination programme reach. Reporting completeness also varied across countries and indicators, with greater challenges for older adult and HCW reporting than for total population, especially in 2024. Further, estimates were based on country-reported administrative data and are therefore subject to reporting artefacts. Comparisons between 2021-2023 and 2024 should also be interpreted cautiously because the monitoring framework and reported indicators changed between periods^21^.

Second, limitations arise from the analytic decisions required to harmonize and summarize heterogeneous country-reported data. For both analytic periods, carrying the latest available report forward treated the last reported value as the best available end-period estimate, which may underestimate vaccination activity where additional doses were administered but not subsequently reported to WHO. In addition, coverage estimates were capped at 100% to avoid implausible values, although values requiring capping may reflect numerator-denominator mismatch, reporting artefacts, or differences between reported target groups and standard population estimates. Further, the 2021-2023 HCW numerator adjustment used total population coverage as a pragmatic proxy to estimate vaccination among HCWs outside reported HCW vaccination targets, which may not fully reflect HCW-specific uptake. These analytic choices supported comparability across countries, indicators, groups, and periods, but should be considered when interpreting specific estimates.

Despite these limitations, the broad temporal patterns and between-country disparities were consistent and sufficiently marked to support the main conclusions. The findings also highlight the need to strengthen immunization information systems for adult and priority group vaccination. Recent WHO guidance on monitoring COVID-19 vaccination coverage, including alongside seasonal influenza and respiratory syncytial virus (RSV) vaccination, provides practical support for countries strengthening routine monitoring of adult and priority group vaccination programmes^33^.

Overall, this global analysis shows that many countries achieved substantial primary series COVID-19 vaccination coverage during the emergency period. These achievements, however, were marked by substantial and persistent disparities across regions and country income groups, reflecting inequities in both vaccine access and roll-out capacity. As programmes moved beyond emergency deployment, the central challenge shifted toward sustaining booster dose vaccination and periodic revaccination of priority groups. Lower booster dose coverage and low, heterogeneous uptake in 2024 suggest that this transition remains incomplete. Together, these findings point to the need for equitable access to safe and effective vaccines, sustainable financing for adult immunization, stronger delivery platforms, fit-for-purpose monitoring systems, and operational capacity to deploy vaccines rapidly and repeatedly at scale. These capacities will be needed both to enhance life-course vaccination benefits and to support future outbreak, epidemic, and pandemic vaccine responses.

## Data availability

The country-reported COVID-19 vaccination data and derived analysis datasets supporting this study are available in the project repository at https://github.com/brooksdonald/global-covid19-vaccination and in the archived release on Zenodo at https://doi.org/10.5281/zenodo.21076717. These datasets reflect WHO COVID-19 vaccination reporting records as extracted on 30 June 2026; subsequent updates are not reflected. Reuse of underlying data values remains subject to the terms of the original data providers, as described in the repository data terms notice.

## Code availability

The analysis code used to clean, process, analyze, and visualize the data is available in the project repository at https://github.com/brooksdonald/global-covid19-vaccination and in the archived release on Zenodo at https://doi.org/10.5281/zenodo.21076717. The code includes scripts used to generate the analytic datasets, summary estimates, figures, and supplementary outputs reported in this study. The analysis code is licensed under the MIT license.

## Acknowledgements

We gratefully acknowledge Rafael Panlilio (CoVDP/UNICEF), Alex Karari (CoVDP/Dalberg), Soha Hourani (WHO HQ), Sebastian Dodt (Dalberg), Denis Sementsov (Dalberg), Jeremy Cooper (Gates Foundation), Kelli Cappelier (CoVDP/JSI), Randie Gibson (WHO HQ), Claudia Steulet (WHO HQ), Theodore Kalmoumenos (WHO EURO), Sheillah Nsasiirwe (WHO AFRO), Boris Pavlin (WHO HQ), and Zyleen Kassamali (WHO HQ) for their contributions to COVID-19 vaccination monitoring. We also thank ministries of health, health system staff, and WHO country office staff for their essential role in collecting, validating, reporting, and interpreting COVID-19 vaccination data throughout the emergency and early post-emergency periods.

## Author contributions

DJB and AG had full access to all data in the study and take responsibility for the integrity of the data and the accuracy of the analysis.

DJB, AG, LC, MGD, and KLO conceived the study. DJB led the study design, data curation, analysis, visualization, and drafting of the manuscript. DJB, AG, LD, YN, CS, FM, AAt, MC, MVG, RL, ET, RP, JK, GG, and BB contributed to data extraction, validation, and interpretation. LC, AAç, FM, RHD, MC, MVG, RL, RP, OM, GG, DCB, SVD, AWS, MGD, MVK, and KLO contributed to methodological review and interpretation of findings. DJB, FM, MV, ET, RP, OM, GG, DCB, AWS, MGD, and KLO provided technical and strategic input on COVID-19 vaccination monitoring and policy interpretation. All authors critically reviewed the manuscript, approved the final version, and agree to be accountable for the work.

