## Supplementary Information File for "Global COVID-19 vaccination uptake across the emergency and early post-emergency periods, 2021-2024"

### Supplementary Methods

#### Supplementary Methods S1. Reporting mechanisms and data compilation

COVID-19 vaccination data were reported to the World Health Organization (WHO) through global and regional reporting mechanisms. Reporting pathways varied across regions and over time, reflecting differences in regional systems, national reporting arrangements, and the transition from emergency COVID-19 vaccination monitoring to routine immunization monitoring.

For 2021-2023, cumulative total population vaccination data were collected through WHO regional reporting processes. Data sources included direct country reporting through digital platforms, submissions to WHO country and regional offices by email, and extraction from Member State health ministry portals and websites. These data were requested and compiled weekly at the global level, with some regional variation in reporting frequency.

Cumulative vaccination data for older adults and health and care workers (HCWs) were collected through the WHO-UNICEF electronic Joint Reporting Form (eJRF) COVID-19 module and WHO regional reporting processes. These data were requested and compiled monthly at the global level, with some regional variation in reporting frequency. Data submitted through the eJRF were reviewed by WHO staff, with follow-up to countries where needed to clarify reported values to improve data quality.

For 2024, end-year COVID-19 vaccination data were collected primarily through the annual 2024 routine immunization eJRF exercise. For most European Union Member States, end-2024 data were obtained through the European Surveillance System. Quarterly COVID-19 vaccination data were also requested and compiled during the first three quarters of 2024 through the eJRF COVID-19 module and WHO regional reporting processes, with some regional variation in reporting frequency. These quarterly data were used where end-year reports were unavailable or to support reconciliation of annual uptake values.

**Supplementary Methods S2. Anomaly detection and correction of cumulative vaccination time series**

The anomaly detection and correction framework was applied to reported cumulative COVID-19 vaccination time series for 2021-2023 before descriptive analysis. Weekly and monthly reported data were organized on a common monthly analytic time scale. Cleaning was applied to cumulative series for total doses administered, at least one dose, complete primary series (CPS), and booster doses in the total population, and to cumulative counts for at least one dose, CPS, and booster doses among older adults and HCWs. Cleaning was performed separately within each country and indicator.

For older adult and HCW target group data during 2021-2023, analytic records were restricted to reports with non-missing and non-zero CPS counts, which were used to identify valid reporting rows. When more than one target group record was available for the same country and reporting month, the record with the largest CPS count was retained. The corresponding at least one dose, CPS, and booster dose values from that selected row were then used in subsequent cleaning and coverage analyses.

The first stage identified structural inconsistencies in cumulative time series, particularly downward steps between successive non-missing observations. For each country-indicator series, observations were ordered by time and screened recursively for violations of monotonicity. When a downward step was identified, the surrounding non-missing observations were examined to determine whether the irregularity was more consistent with an implausibly high earlier value or an implausibly low later value. Consecutive observations identified as belonging to the same irregular sequence could be removed together. Observations identified as implausible were set to missing.

In the second stage, internal missing values, including gaps in reporting and gaps created by anomaly removal, were replaced by linear interpolation between the surrounding non-missing cumulative values. Interpolated values were rounded to integers and constrained to preserve monotonicity after interpolation and rounding. Following the final available observation, the latest cumulative value was carried forward through the end of the analysis period.

A further interpolation step was used to construct trajectories before the first reported cumulative value. For total doses administered and at least one dose indicators in the total population, older adults, and HCWs, values were interpolated linearly from zero at the country-specific date of vaccine programme introduction to the first non-missing reported value. For CPS indicators, interpolation began two months after vaccine introduction, reflecting the expected interval between vaccine programme introduction and completion of the primary series. Because analyses were conducted on a monthly time scale, programme introduction dates and reporting months were represented by the 15th day of the corresponding month for interpolation.

Total population booster series were retained from the first reported value without interpolation of preceding months. For older adult and HCW booster indicators, the country's first reported total population booster month was used as a proxy for the start of booster vaccination. Values were interpolated linearly from zero in that month to the first non-missing population-group-specific booster value. No pre-report interpolation was applied when a total population booster value was unavailable before the first population group-specific report or when both were first reported in the same month. Outside these intervals, the monotonicity-cleaned series were retained unchanged.

After cleaning and interpolation, cumulative counts of vaccinated persons were capped at the relevant population denominator to prevent coverage estimates from exceeding 100%. Total doses administered were not capped because individuals could receive multiple doses.

This implementation was intended to correct structural artefacts and construct consistent monthly trajectories between observed cumulative reports. Interpolation assumed a linear increase between specified starting points and first reports or between surrounding reported values, while carry-forward after the final report assumed no additional reported vaccination activity. The procedure did not otherwise extrapolate vaccination activity beyond the observed reporting period. As noted in the main text, the framework was not applied to 2024 annual uptake values. A fuller methodological evaluation of the anomaly detection framework, including its performance characteristics and applicability beyond this analysis, is outside the scope of the present manuscript and will be reported separately.

**Supplementary Methods S3. Reporting completeness over time**

To describe reporting completeness during the emergency period, we conducted a descriptive analysis of country reporting using country-reported COVID-19 vaccination data from 1 January 2021 to 31 December 2023. Analyses were restricted to WHO Member States. Reporting was assessed separately for the total population, older adults, and HCWs, and for CPS and booster dose indicators.

Reporting data were extracted from weekly total population reporting files and monthly target group reporting files and organized on a common monthly analytic time scale. For total population reporting, when multiple weekly records were available for the same country-month, the latest report within that month was retained. For older adult and HCW target group reporting, when multiple records were available for the same country-month and target group, the record with the largest CPS count was retained.

Two complementary dimensions of reporting were summarized. First, month-specific reporting was defined as the number of Member States reporting a non-missing and non-zero value for the relevant population group and indicator in a given month. For month-specific reporting counts, countries reporting booster doses were subtracted from CPS reporting counts so that CPS counts represented reporting without concurrent booster reporting.

Second, cumulative reporting completeness was assessed by constructing a complete country-month panel, carrying reported cumulative values forward within countries across subsequent months, and counting the number of Member States with a non-missing value for each population group and indicator. Values of zero were treated as missing for this analysis. A small number of identified reporting artefacts in cumulative reporting series were set to missing before carry-forward. Cumulative reporting completeness was summarized as counts of reporting Member States and, for income group summaries, as the proportion of Member States in each income group with a reported value.

### Supplementary Tables

**Supplementary Table S1. Operational definitions of WHO-requested COVID-19 vaccination uptake/coverage indicators used in this study (2021-2024)**

| Period | Indicator | Operational definition | Numerator | Denominator |
|---|---|---|---|---|
| 2021-2023 | Total doses administered | Cumulative number of COVID-19 vaccine doses administered (all dose numbers combined) in the reporting population. | Total number of doses administered (all products; all dose numbers). | Total population (if expressed per 100) or none (if absolute counts). |
| 2021-2023 | At least one dose coverage | Proportion of the relevant population that has received ≥1 dose of a COVID-19 vaccine (cumulative). | Number of people who received ≥1 dose. | Size of the relevant population group. |
| 2021-2023 | Complete primary series coverage | Proportion of the relevant population that has received the last recommended dose to complete the primary schedule for the vaccine product used. | Number of people who received the last recommended primary-series dose (product-specific). | Size of the relevant population group. |
| 2021-2023 | At least one booster dose coverage | Proportion of the relevant population that has received ≥1 booster dose (an additional dose administered after completion of a primary series, per national policy during 2021-2023). | Number of people who received ≥1 booster dose. | Size of the relevant population group. |
| 2024 | Individuals vaccinated with a COVID-19 | Number (and corresponding uptake %) of individuals receiving one COVID-19 | Number of individuals vaccinated with one dose during | Total number of individuals in the corresponding |

| | | | | |
|---|---|---|---|---|
| | vaccine dose | vaccine dose during the calendar year, regardless of whether it is primary vaccination or periodic revaccination. | the year, optionally disaggregated by population group. | population group (for uptake %). |

**Supplementary Table S2. WHO-recommended priority population group definitions for COVID-19 vaccination monitoring and reporting**

| Population group | WHO-recommended definition / guidance |
|---|---|
| Older adults | Minimum age threshold to be defined by countries; commonly ≥50 or ≥60 years. |
| Health and care workers | Definition to be defined by countries; WHO recommends using the “Health professional” definition aligned with the International Classification of Health Workers. |
| Adults | Adult age range to be defined by countries; commonly 18–49 or 18–59 years. |
| Children and adolescents | Age range to be defined by countries; commonly 5–17 years. |
| Adults with chronic conditions / Individuals with chronic conditions | May include persons with significant comorbidities (examples provided): diabetes; chronic lung disease; heart, liver, kidney disease; severe obesity (BMI >40); immunocompromised (e.g., active cancer, transplant recipients, immunodeficiency on immunosuppressives, and specific HIV-related criteria). |
| Pregnant women (pregnant persons) | The questionnaire uses “pregnant women,” while noting that transgender men and other gender-diverse people can also become pregnant; vaccination of all pregnant persons may be reported under this category. |

**Supplementary Table S3. Maximum average daily COVID-19 vaccination rate and timing of peak activity during 2021-2023, by income group and region**

Maximum values correspond to the peak of the trailing 28-day mean of daily vaccination rates, expressed as a percentage of the total population. Dates indicate when the maximum average daily vaccination rate was reached for each income group and region.

| Category | Date of peak | Peak DVR |
|---|---|---|
| **All** | | |
| Global | 2021-07-11 | 0.48% |
| **Income group** | | |
| HIC | 2021-07-10 | 0.64% |
| UMIC | 2021-07-11 | 0.82% |
| LMIC | 2022-01-23 | 0.44% |
| LIC | 2022-12-18 | 0.12% |
| **WHO region** | | |
| AFR | 2022-07-17 | 0.17% |
| AMR | 2021-11-26 | 0.50% |
| EMR | 2021-09-29 | 0.40% |
| EUR | 2021-07-11 | 0.69% |
| SEAR | 2021-09-28 | 0.56% |
| WPR | 2021-06-27 | 0.95% |
| **COVAX participation category** | | |
| AMC | 2021-12-21 | 0.37% |
| Self-financing | 2021-06-27 | 0.92% |

### Supplementary Figures

**Supplementary Figure S1: Reporting completeness for COVID-19 vaccination data by population group, 2021–2023**

(a) Monthly and cumulative reporting by population group. Bars show the monthly number of WHO Member States submitting coverage data for complete primary series (CPS) only (blue) or for both CPS and at least one booster dose (yellow), shown separately for the total population, older adults, and health and care workers. Overlaid lines show the cumulative number of Member States that had reported CPS data (blue) or booster data (yellow) at least once by each month. The horizontal dashed line indicates the total number of WHO Member States (194).

(b) Cumulative reporting completeness by World Bank income group. Lines show the percentage of Member States in each income group that had reported CPS or booster data at least once by each month, stratified by population group and indicator. End labels indicate the final cumulative reporting percentage at the end of 2023.

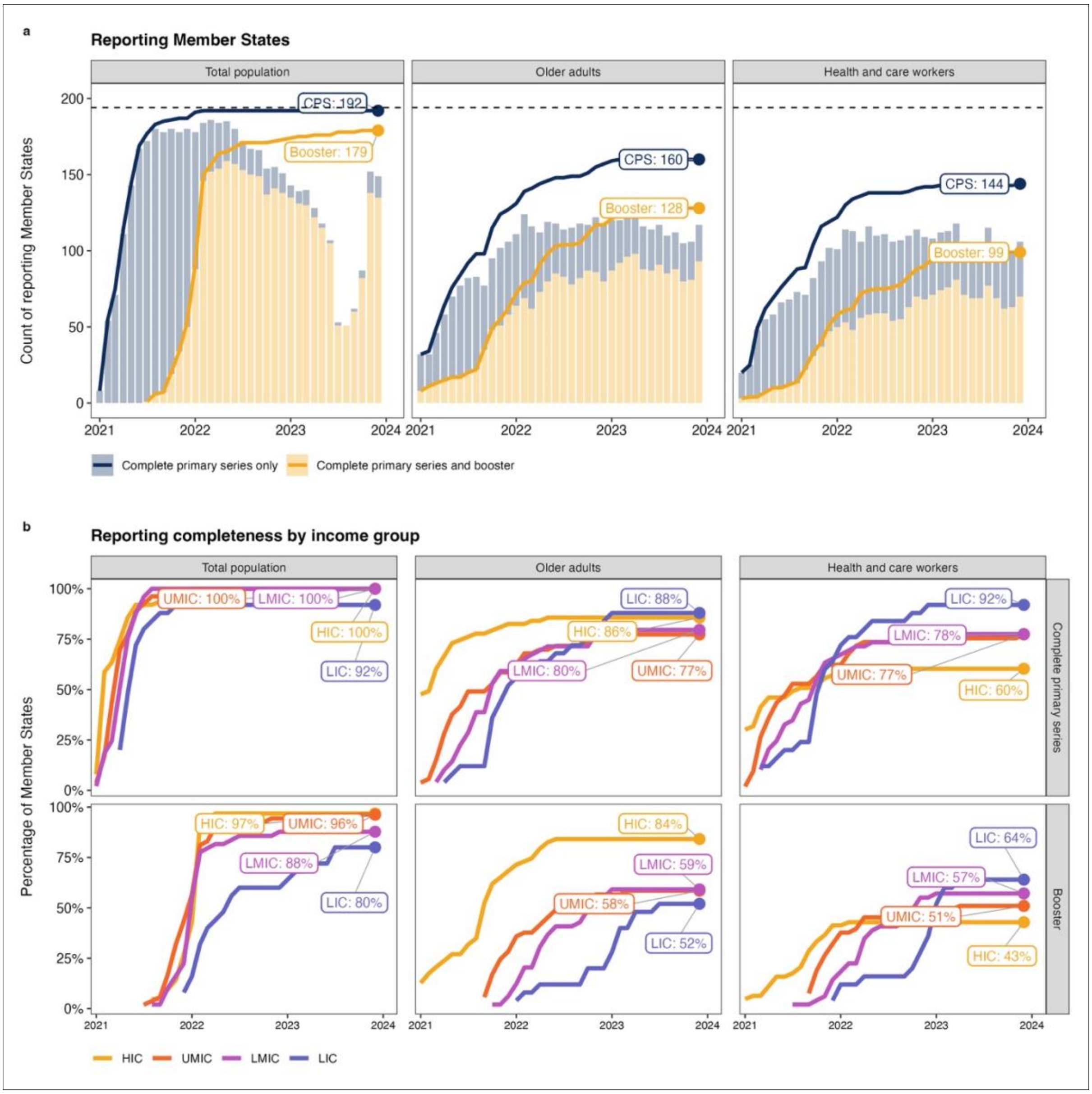


**Supplementary Figure S2: Complete primary series COVID-19 vaccination coverage in priority groups relative to the total population, end-2021 and end-2023**

(A) Country-level complete primary series coverage in the total population (x-axis) versus older adults (top row) and health and care workers (bottom row) (y-axis), at end-2021 and end-2023. Each point represents a country and is colored by World Bank income group. The diagonal reference line indicates parity between priority-population and total-population coverage (points above the line indicate higher coverage in the priority population).

(B) Difference from parity (priority population minus total population, percentage points) by World Bank income group for the same time points and population groups. Distributions are shown as violins with embedded boxplots (median and interquartile range).

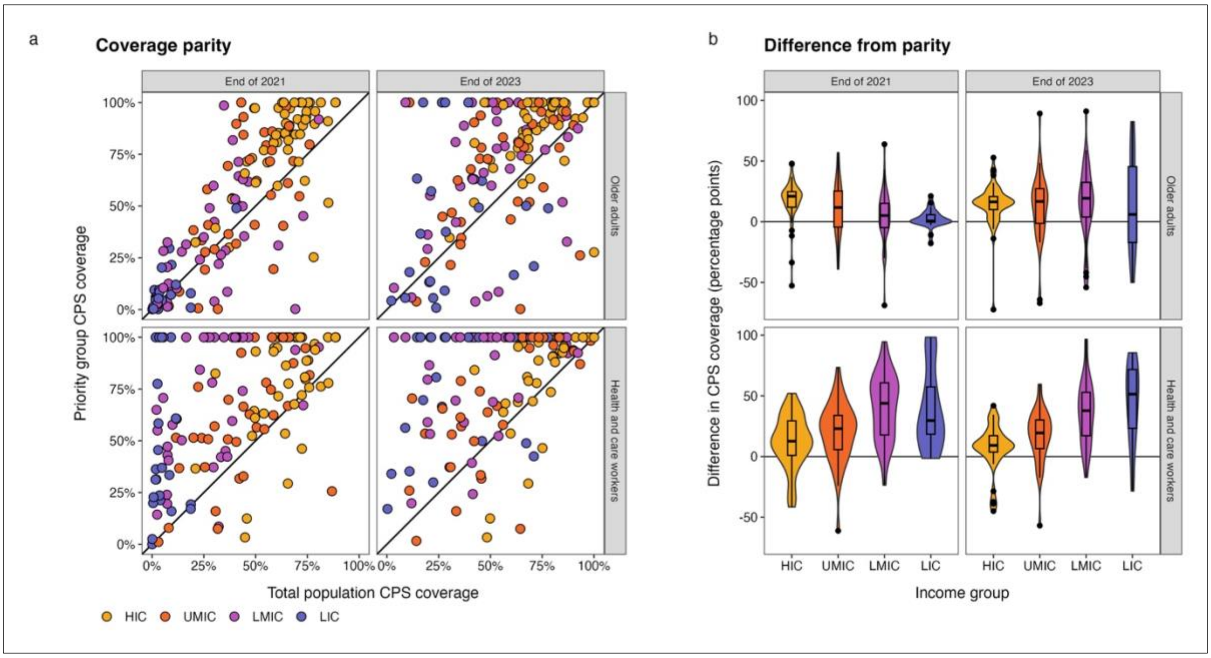


**Supplementary Figure S3: COVID-19 vaccination coverage by population group and World Bank income group, 2021-2023**

Panels show the temporal evolution of population-weighted average vaccination coverage (%) for the total population, older adults, and health and care workers, stratified by World Bank income group. Lines represent coverage with at least one dose, complete primary series, and at least one booster dose, calculated from reported cumulative uptake as a percentage of the relevant population denominator and aggregated within each income group using population weights. Values shown at the end of each line indicate the end-2023 coverage level for that series.

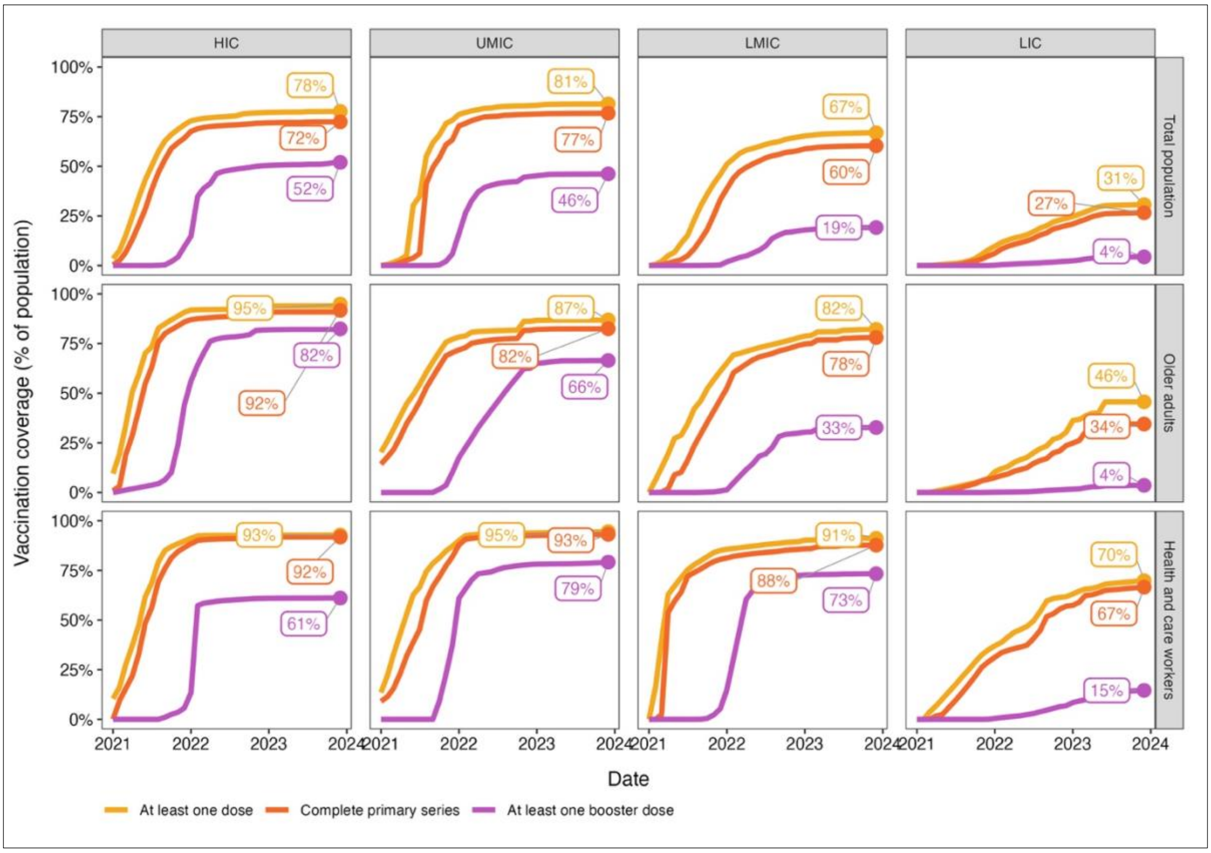


**Supplementary Figure S4: COVID-19 vaccination coverage (median, IQR) by population group and World Bank income group, 2021-2023**

Panels show the distribution of country-reported vaccination coverage over time for the total population, older adults, and health and care workers, stratified by World Bank income group. For each month, the median coverage across reporting countries is shown for complete primary series and at least one booster dose, with shaded bands indicating the interquartile range (IQR; 25th-75th percentiles). Coverage is expressed as a percentage of the relevant population denominator and summarizes heterogeneity in uptake within each income group across the study period. Monthly estimates are shown only when reporting countries represented at least 10% of countries that reported data for the relevant population group and vaccination indicator during 2021-2023.

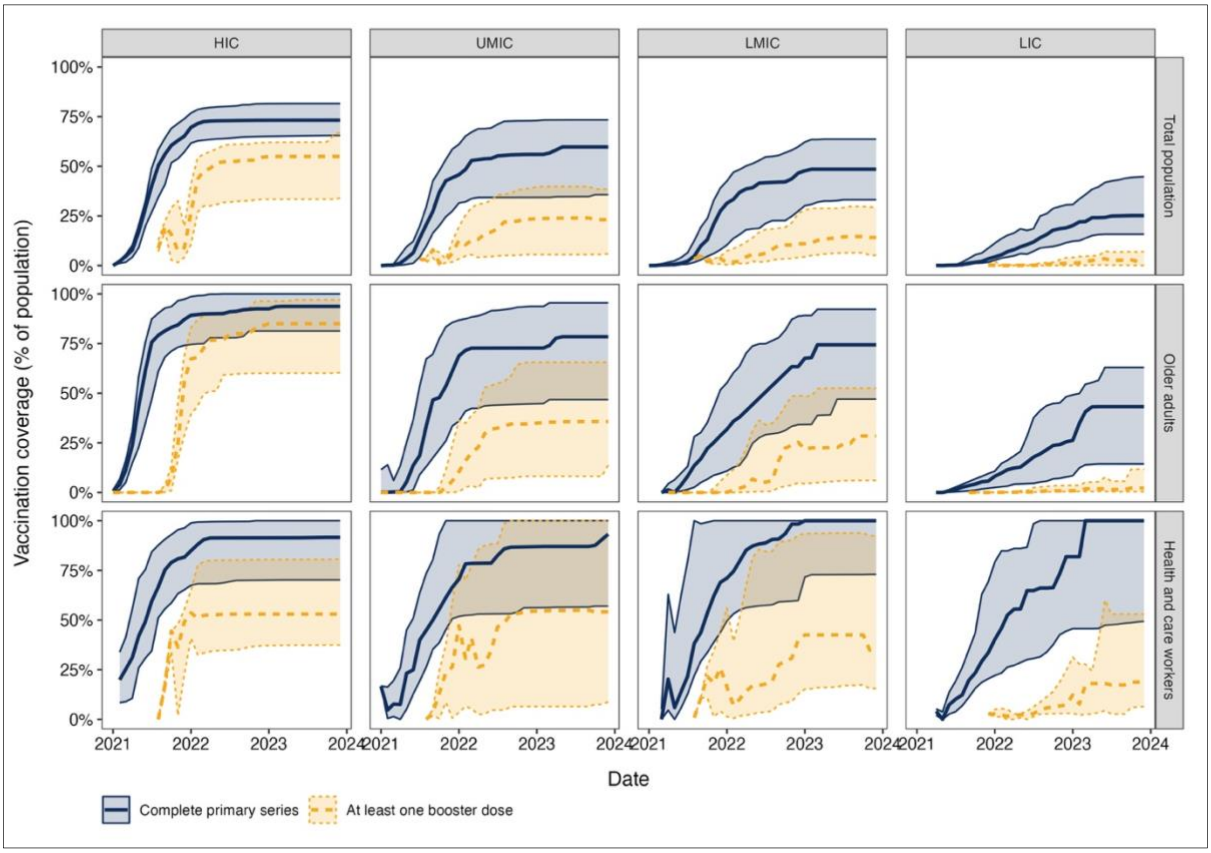


**Supplementary Figure S5: Weighted average daily COVID-19 vaccination rate in the total population, by WHO region and World Bank income group, 2021-2023**

Lines show the population-weighted average of country-level trailing 28-day mean daily vaccination rates, expressed as the percentage of the total population vaccinated per day. Estimates are shown by WHO region (top panel), World Bank income group (middle panel), and global (bottom panel). Dots mark the peak value for each series.

194

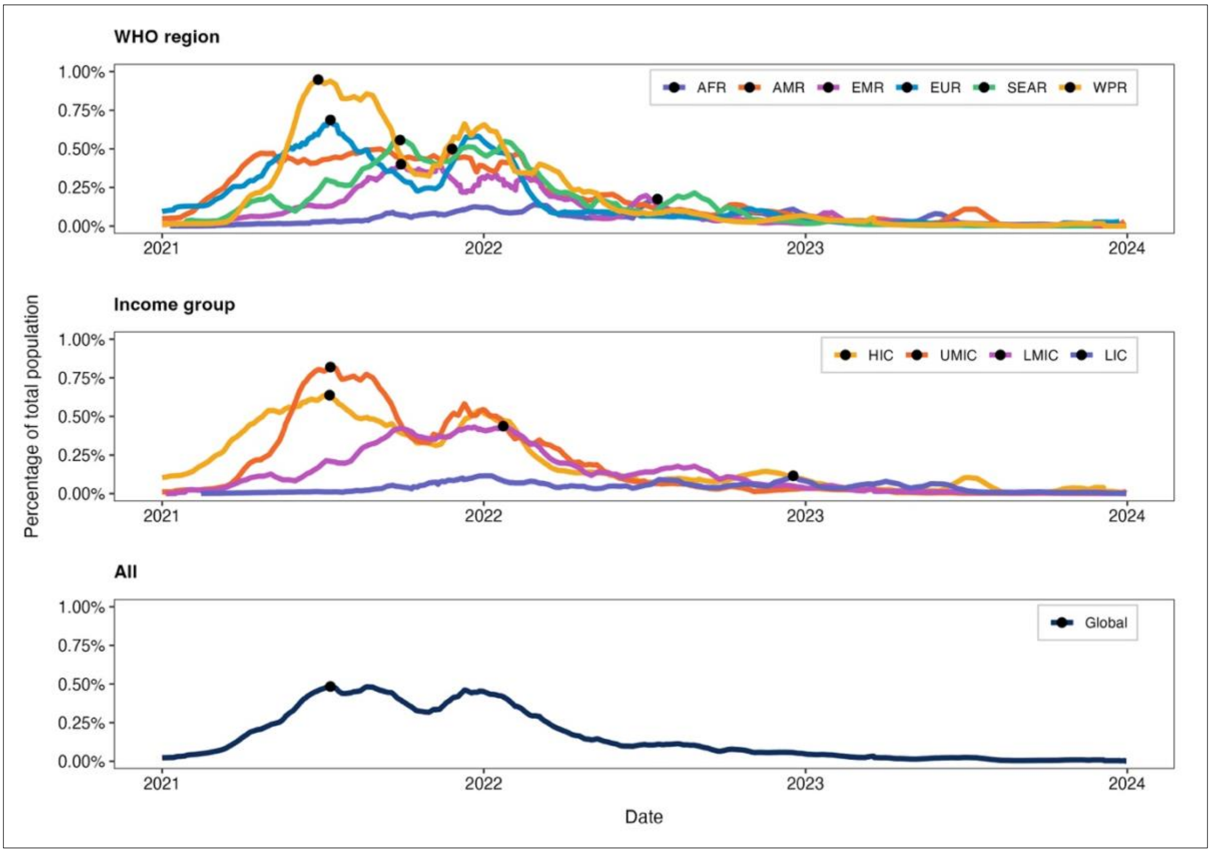

WHO region
AFR AMR EMR EUR SEAR WPR
1.00%
0.75%
0.50%
0.25%
0.00%
2021
2022
2023
2024
Income group
HIC UMIC LMIC LIC
Percentage of total population
All
Global
Date
